# Global burden of gout in the working-age population (15–64 years), 1990–2021: a systematic analysis of the Global Burden of Disease Study 2021

**DOI:** 10.64898/2026.01.30.26345202

**Authors:** JF Zhang, I-Han Cheng, James Cheng-Chung

## Abstract

Gout is the most common inflammatory arthritis worldwide, traditionally associated with older males (2024c, 2025). However, many studies focus on the elderly, leaving a gap in understanding the burden of gout among working-age individuals. We aimed to quantify the global burden of gout in the working-age population (WAP, 15–64 years) and characterise trends, inequalities, and future projections, using Global Burden of Disease (GBD) 2021 data.

We extracted GBD 2021 estimates for gout in 204 countries and territories from 1990 to 2021 for ages 15–64. Disease burden was assessed through incidence, prevalence, and years lived with disability (YLDs), along with corresponding age-standardised rates (per 100,000). Trend analysis included calculating the estimated annual percentage change (EAPC) of age-standardised rates and using join point regression to detect inflexion points. We quantified contributions of demographic drivers (population growth and ageing) versus epidemiologic changes in gout rates using decomposition analysis. Cross-national inequalities were evaluated with the Slope Index of Inequality (SII) and concentration index for gout metrics across the socio-demographic spectrum. A forecasting model (based on GBD data and a mixed-effects temporal model) projected gout burden in WAP through 2045.

Globally in 2021, there were 6.08 million (95% UI: ∼5.5–6.6) new gout cases and 32.7 million (UI: ∼30–35) prevalent gout cases among people aged 15–64 years. The age-standardised incidence rate (ASIR) was approximately 105 per 100,000, and the age-standardised prevalence rate (ASPR) was about 510 per 100,000 in 2021. These rates rose modestly since 1990 (EAPCs ∼0.5–0.8%/year), reflecting a gradual upward trend. No major joinpoints were indicated to have sudden trend shifts over the 1990–2021 period. Males in the working-age group had about 3-fold higher gout prevalence than females, mirroring known sex disparities. High-income regions (e.g., Australasia and North America) exhibited the highest WAP gout rates. In contrast, some low-income areas (e.g., parts of sub-Saharan Africa and Latin America) had the lowest. A moderate positive correlation between national gout burden and Socio-demographic Index (SDI) was observed. Health disparities widened over time: the gout concentration index increased, indicating the disease became more unequally distributed, disproportionately affecting higher-SDI populations by 2021. From 1990 to 2021, the total number of WAP gout cases increased substantially. Decomposition analysis revealed that population growth accounted for ∼54.4% of this increase, with population ageing and rising age-specific gout prevalence contributing ∼20% and ∼25%, respectively (Table 1). We project that by 2045, the number of working-age gout cases will rise to ∼45 million, a 40% increase from 2021. Despite growing absolute cases, age-standardised rates are expected to stabilise or increase only slightly in the coming decades, assuming current trends hold.

The burden of gout in working-age adults has increased globally over the past three decades and is expected to continue growing due to demographic expansion. This first comprehensive analysis of the working-age population reveals that gout is not merely a disease of the elderly – a majority of global cases occur before the age of 65, with significant implications for productivity and healthcare systems. Urgent, age-targeted prevention strategies (e.g. obesity and metabolic syndrome management) and improved access to care for younger gout patients are warranted to curb the rising burden and reduce future disability.

## Introduction

Gout is a metabolic inflammatory arthritis caused by the deposition of monosodium urate crystals in joints due to chronic hyperuricaemia. (Dalbeth et al., 2021a). Clinically, it manifests in episodic flares of severe joint pain and swelling and can progress to chronic tophaceous arthropathy if untreated. Gout is the most common form of inflammatory arthritis worldwide (2024c, Danve and Neogi, 2020)., with an estimated 41.2 million adults affected by gout worldwide in 2017(Danve and Neogi, 2020). The prevalence of gout varies widely, ranging from under 1% to about 6–7% of the population in different countries(Dehlin et al., 2020). Both the prevalence and incidence of gout have been rising across the globe over the past few decades(Dehlin et al., 2020). In 2017, the global prevalence of gout was roughly double that of 1990 (increasing from 20.2 million to 41.2 million cases)(Danve and Neogi, 2020), reflecting substantial growth in disease burden.

Gout primarily affects adult men and older individuals. Men have a significantly higher gout prevalence than women. Globally, gout is about three times more common in males than in females. The risk of gout increases with age, peaking in older age groups (Dehlin et al., 2020). But a considerable burden of disease also occurs in middle-aged populations. The geographic distribution of gout is uneven; the highest prevalence rates are observed in specific high-risk populations and regions (for example, some Pacific Island nations report gout in up to ∼6–7% of adults)(Dehlin et al., 2020). Historically, developed (high-income) countries have had a higher gout burden than developing countries(Dehlin et al., 2020). Nevertheless, all regions of the world have experienced a rise in gout prevalence and incidence in recent years. Notably, such areas as high-income North America and Australasia currently exhibit the most significant prevalence of gout. At the same time, some of the sharpest relative increases over time have also been reported in these regions and others.

Multiple modifiable risk factors underlie the development of gout. Many factors that contribute to elevated serum urate, including purine-rich diets, excessive alcohol intake, obesity, metabolic syndrome, chronic kidney disease, and the use of certain medications (e.g. diuretics), are known to increase the risk of gout (Dalbeth et al., 2021b). In particular, the global rise in obesity and related comorbidities (such as type 2 diabetes and hypertension) has been identified as a major driver of the increasing prevalence of gout (Dehlin et al., 2020). High body mass index and other components of the metabolic syndrome promote hyperuricaemia and gout, and these factors are increasingly common worldwide (Dehlin et al., 2020). Chronic kidney disease is another significant contributor, as impaired renal urate excretion can lead to hyperuricaemia (Dalbeth et al., 2021b). The strong associations between gout, obesity, and kidney dysfunction mean that trends in these conditions are closely intertwined (Dehlin et al., 2020).

Gout flares cause intense pain and disability, which can significantly impact patients’ daily activities and work productivity. Because gout often onsets in mid-life and affects individuals during their working years, it has substantial socioeconomic consequences. Patients with gout experience higher rates of work absenteeism; for example, employees with gout have been shown to take approximately 4–5 more sick days per year on average compared to those without gout (Kleinman et al., 2007). Recurrent gout attacks and chronic arthropathy can thus impose a considerable burden on both the healthcare system and the workforce. This global burden of gout in the working-age population warrants heightened attention, as effective interventions to control risk factors and improve gout management could yield significant benefits in reducing disability and improving productivity.

## Methods

### Data Source

We conducted a population-level trend analysis using data from the Global Burden of Disease (GBD) 2021 study. The GBD2021 (https://vizhub.healthdata.org/gbd-results/), provides a comprehensive assessment of mortality and disability related to hundreds of diseases, injuries, and risk factors across 204 countries and territories from 1990 to 2021(2024b). For the current analysis, data on gout were derived from this database. The GBD study categorises countries and regions into seven super areas, which are further subdivided into 21 GBD regions, based on geographical location.(2024b). Additionally, the Socio-demographic Index (SDI) is employed as a composite indicator of a country or region’s socio-economic development level. According to the regional economic development level, the GBD study classifies countries and regions into five categories: high SDI, high-middle SDI, middle SDI, middle-low SDI, and low SDI regions.(2024a). gout in GBD is defined according to standard diagnostic criteria -the M10 code from version 10 of the International Classification of Diseases (ICD) and the corresponding ICD-9-CM diagnosis code (274: Gout) to identify cases of gout in medical claims data.

### Study design and data source

For this analysis, we extracted GBD 2021 results for gout specifically in the working-age population (WAP), defined as ages 15 to 64 years.

#### Measures of disease burden

Key measures analyzed in this study included both absolute burden (case counts) and rate measures for gout among ages 15–64. We analysed incidence (new cases per year), prevalence (total number of individuals living with gout), and years lived with disability (YLDs), as well as crude rates (per 100,000 population), which reflect the cumulative health loss due to gout-related disability. Since gout causes little to no direct mortality, we did not evaluate disease-specific mortality or disability-adjusted life years (DALYs); for this condition, DALYs would be nearly equivalent to years lived with disability (YLDs).

For each of these measures, we examined both crude rates (expressed per 100,000 population) and age-standardised rates to allow comparisons across populations and over time. For each age group, as well as the age-standardised incidence rate (ASIR), age-standardised prevalence rate (ASPR), and age-standardised YLD rate (ASYR), were calculated using the formula ∑(a_i_ × w_i_)/∑w_i_, where a_i_ denotes the age-specific rate in the i-th age group and w_i_ represents the corresponding proportion in the World Health Organization (WHO) standard population. The uncertainty of estimates was quantified by generating 95% uncertainty intervals (UIs) through 1,000 posterior simulations, with statistical significance defined as a 95% UI excluding zero. All statistical analyses were performed using R software (version 4.4.0).

#### Analytical approaches

We employed several analytical techniques to characterise trends and disparities in the burden of gout among the working-age population from 1990 to 2021., The temporal trends of age-standardised rates were assessed using the estimated annual percentage change (EAPC) (Hankey et al., 2000). An EAPC > 0 indicates an increasing trend, and < 0 a decreasing trend; it was considered statistically significant if the confidence interval (CI) did not include 0. We also examined the absolute change in case numbers and rates between 1990 and 2021. In addition to EAPC, Joinpoint regression (version 5.2.0; National Cancer Institute, Rockville, MD, USA) analysis was employed to identify significant changes in the temporal trends of age-standardised rates. This method tests whether a multi-segment linear model fits the data better than a single linear trend and allows the identification of time points (“joinpoints”) at which the trend significantly changes(Kim et al., 2000).

Moreover, to evaluate the health inequality between population subgroups stratified by socioeconomic, demographic, geographic, or other socially relevant factors, we calculated the absolute and relative inequality metrics(Hosseinpoor et al., 2023), and decomposition analysis was conducted to partition observed variations in incidence, prevalence, and YLDs into population growth, aging, and epidemiological transitions. A detailed description of the methods can be found in the Supplementary Material.

## Results

### Global burden in 2021

Compared with 1990, both the incidence and YLDs due to gout in the WAP more than doubled by 2021, while prevalence roughly tripled. In 2021, an estimated 6.08 million new cases (95% uncertainty interval [UI] 3.61 to 9.12 million) occurred, corresponding to an ASIR of 113.20 (95% UI 67.31 to 169.82) per 100,000. Prevalent cases reached 32.74 million (95% UI 21.27 to 46.60 million), yielding an ASPR of 605.03 (95% UI 392.58 to 861.86) per 100,000. The YLDs burden was 1.05 million years (95% UI 0.60 to 1.70 million), corresponding to an age-standardised YLDs rate (ASYR) of 19.46 (95% UI 11.12 to 31.51) per 100,000. By contrast, in 1990, there were 2.83 (95% UI 1.68 to 4.28) million incident cases, 14.43 (95% UI 9.20 to 20.75) million prevalent cases, and 0.47 (95% UI 0.26 to 0.75) million YLDs (Table S1, Figure 1). Figure 2 and Figure 3show the incidence, prevalence, and YLDs in general over the period 1990-2021 for regions with different SDI levels, respectively. Joinpoint regression analyses of global trends from 1990 to 2021 revealed significant increases in ASIR (average annual percentage change [AAPC] = 0.45, 95% confidence interval [CI] 0.44 to 0.46), ASPR (AAPC = 0.56, 95% CI 0.55 to 0.57), and ASYR (AAPC = 0.55, 95% CI 0.53 to 0.55). ASYR exhibited significant joinpoints in 1995, 2001, 2009, 2014, and 2019, with annual percent changes (APCs) progressively shifting from 0.63 (1995-2001) to 1.18 (2001-2009), then decelerating through 0.54 (2009-2014) and 0.41 (2014-2019), before transitioning to negative growth (-0.44) during 2019-2021, and the pattern mirrored in ASIR, ASPR, and sex-stratified analyses (Table S1, Figure 2).

**Figure 1.**
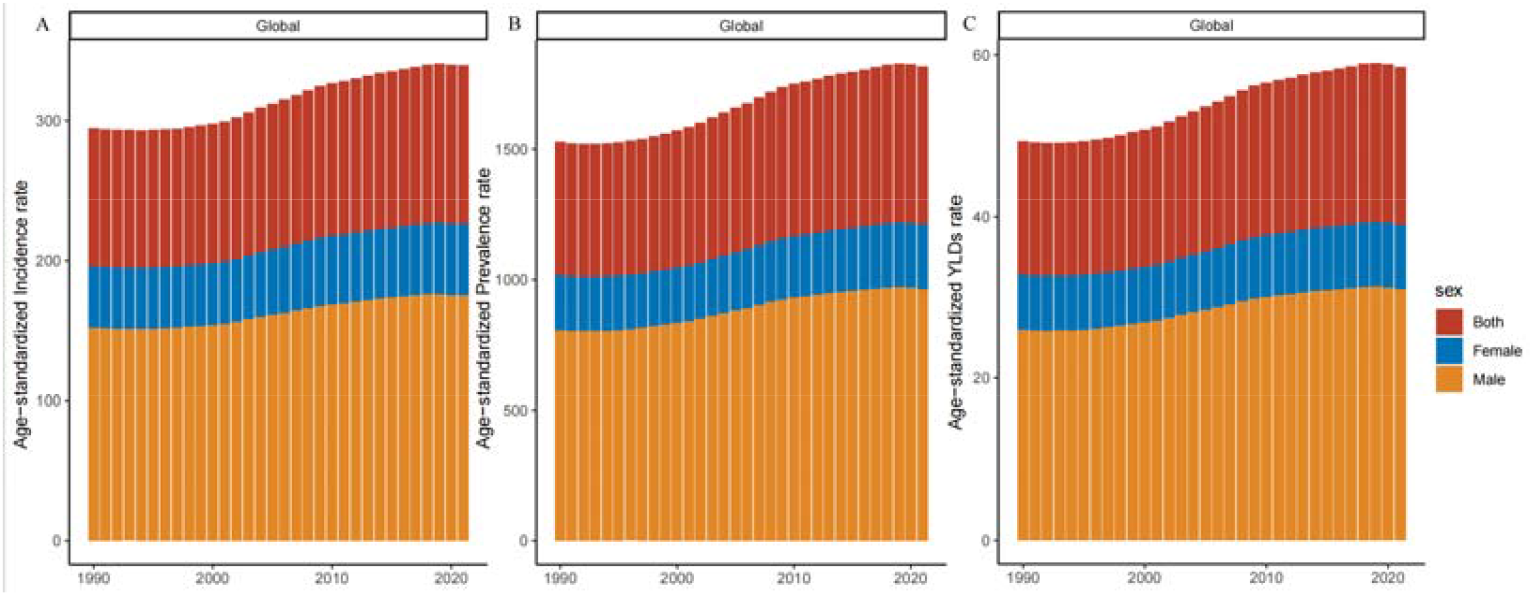
Age-standardised incidence (A), prevalence (B) and YLDs (C) rate of gout from 1990 to 2021. The lengths of the red, blue, and orange bars represent the point estimates for the global, male, and female age-standardised rates.

**Figure 2.**
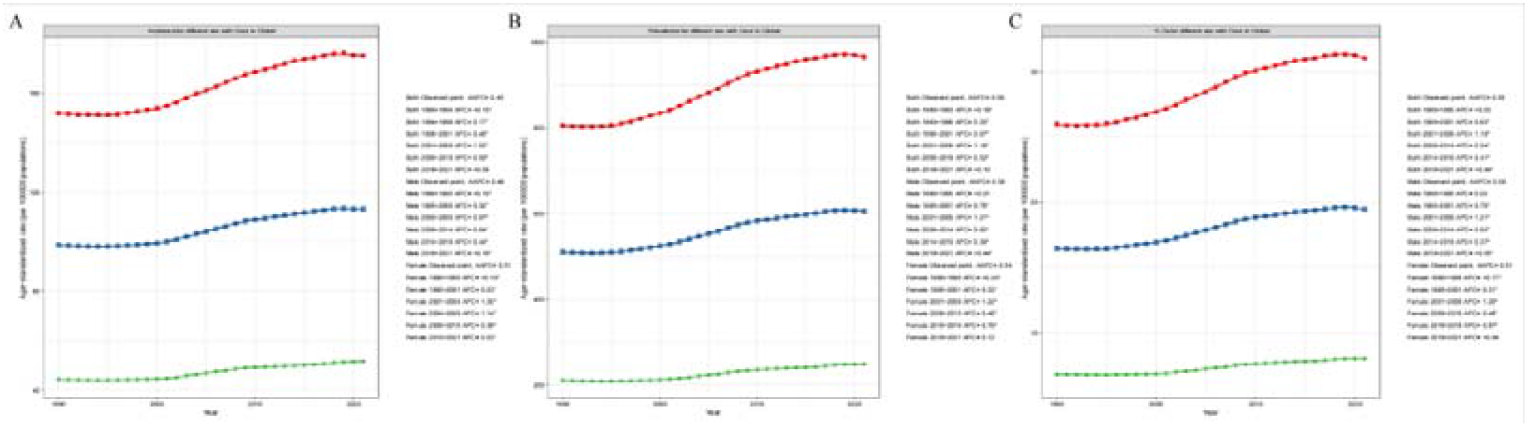
Join point regression analysis of the temporal trends of gout for ASIR (A), ASPR (B), and ASYR (C) from 1990 to 2021. The lines of red, blue, and green represent the ASR of the total, male, and female populations.

**Figure 3.**
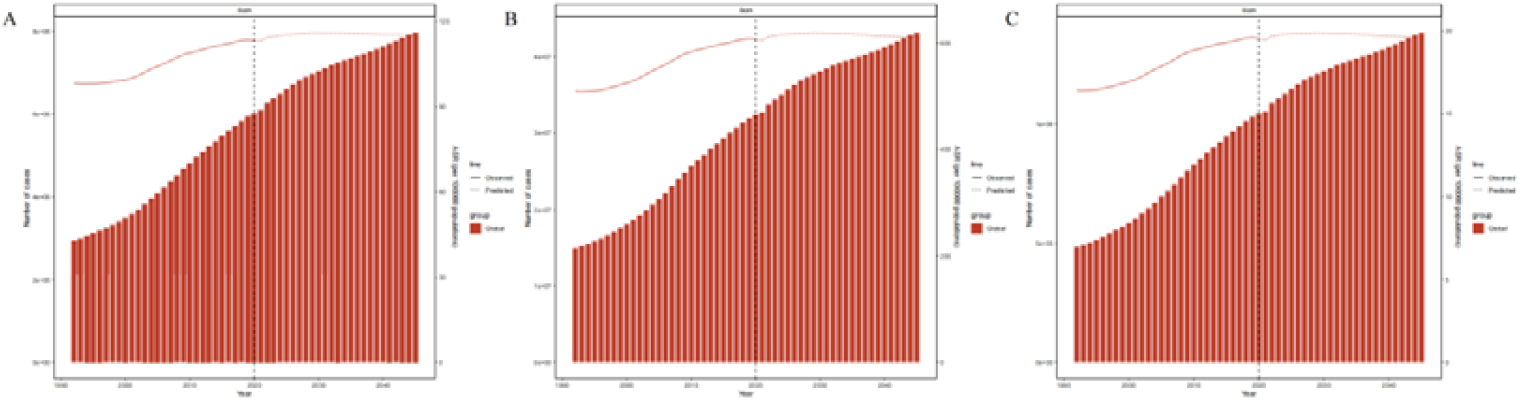
The forecast of the number of cases and age-standardised rates of incidence (A), prevalence (B), and YLDs (C) for gout in 2045. The lines represent age-standardised rates; the bars indicate the number of cases.

Forecasts for 2022 to 2045 indicate a further rise in gout burden among WAP: by 2045, the ASIR is projected to reach 115.03 per 100 000, and both prevalent case numbers and YLDs are expected to increase by approximately one□third—to about 431.63 million cases and 1.38 million YLD years, respectively (Table S2, Figure 3).

### Regional level

In 2021, East Asia (China, Japan, South Korea, North Korea, and Mongolia) reported the most considerable absolute burden among WAP, with 20.44 million incident cases (95% UI, 12.10-30.80 million), 105.06 million prevalent cases (95% UI 66.98 to 151.92 million), and 0.34 million YLDs (95% UI 0.19 to 0.55 million years), accounting for roughly one-third of the global total. Among these five countries, China had the highest ASIR at 161.3 per 100 000 (95% CI 95.6 to 243.4), whereas South Korea exhibited the highest ASPR (818.5 per 100 000, 95% CI 521.1 to 1193.3) and ASYR (26.8 per 100 000, 95% CI 13.9 to 45.3). Nevertheless, East Asia ranked third among the 21 Global Burden of Disease regions in terms of age-standardised incidence, prevalence, and YLDs. High-income North America (United States and Canada) led with an ASIR of 160.0 (95% CI 149.0 to 190.0), an ASPR of 222.0 (95% CI 203.0 to 241.0), and an ASYR of 219.0 (95% CI 200.0 to 238.0) per 100 000 (Table S1, Table S3).

From 1990 to 2021, most regions showed upward trends in the EAPC of incidence, prevalence, and YLD rates; Western Sub-Saharan Africa was the sole exception (EAPCs -0.03 (95% CI, -0.06 to 0) for incidence, -0.02 (95% CI, -0.05 to 0.01) for prevalence, and 0.01 (95% CI, -0.01 to 0.04) for YLDs). Seven regions exceeded the global increase in YLDs: High-income North America (2.19, 95% CI 2.00 to 2.18), Andean Latin America (0.99, 95% CI 0.96 to 1.03), East Asia (0.90, 95% CI 0.81 to 0.98), Australasia (0.82, 95% CI 0.79 to 0.85), Tropical Latin America (0.82, 95% CI 0.79 to 0.85), the Caribbean (0.76, 95% CI 0.74 to 0.78), and Central Latin America (0.74, 95% CI 0.72 to 0.76) (Table S1).

### National level

Driven by population size and SDI, China, the USA, and India had the highest numbers of gout cases and YLDs in 2021, with 10.09 million cases (95% UI, 6.43 to 14.61 million), 4.37 million cases (95% UI 3.09 to 5.86 million), and 3.37 million cases (95% UI 2.12 to 4.94 million), respectively. Collectively, these three countries represented 54.5% of the global gout case burden (Table S3). Across 204 countries, ASIR ranged from 31.6 to 219.0 per 100,000, and ASPR from 179.0 to 1641.1 per 100,000. The highest ASIRs were in New Zealand (219.0, 95% CI 129.7 to 333.0), Canada (202.2, 95% CI 119.8 to 307.9), and Australia (188.8, 95% CI 111.8 to 285.4). The USA (1641.1, 95% CI 1156.5 to 2207.1), Canada (1461.1, 95% CI 974.9 to 2056.3), and Greenland (1407.9, 95% CI 936.8 to 1982.7) had the highest ASPRs, whereas Guatemala had the lowest ASIR (36.6, 95% CI 21.1 to 56.7) and ASPR (179.0, 95% CI 110.9 to 268.5) (Figure 4).

**Figure 4.**
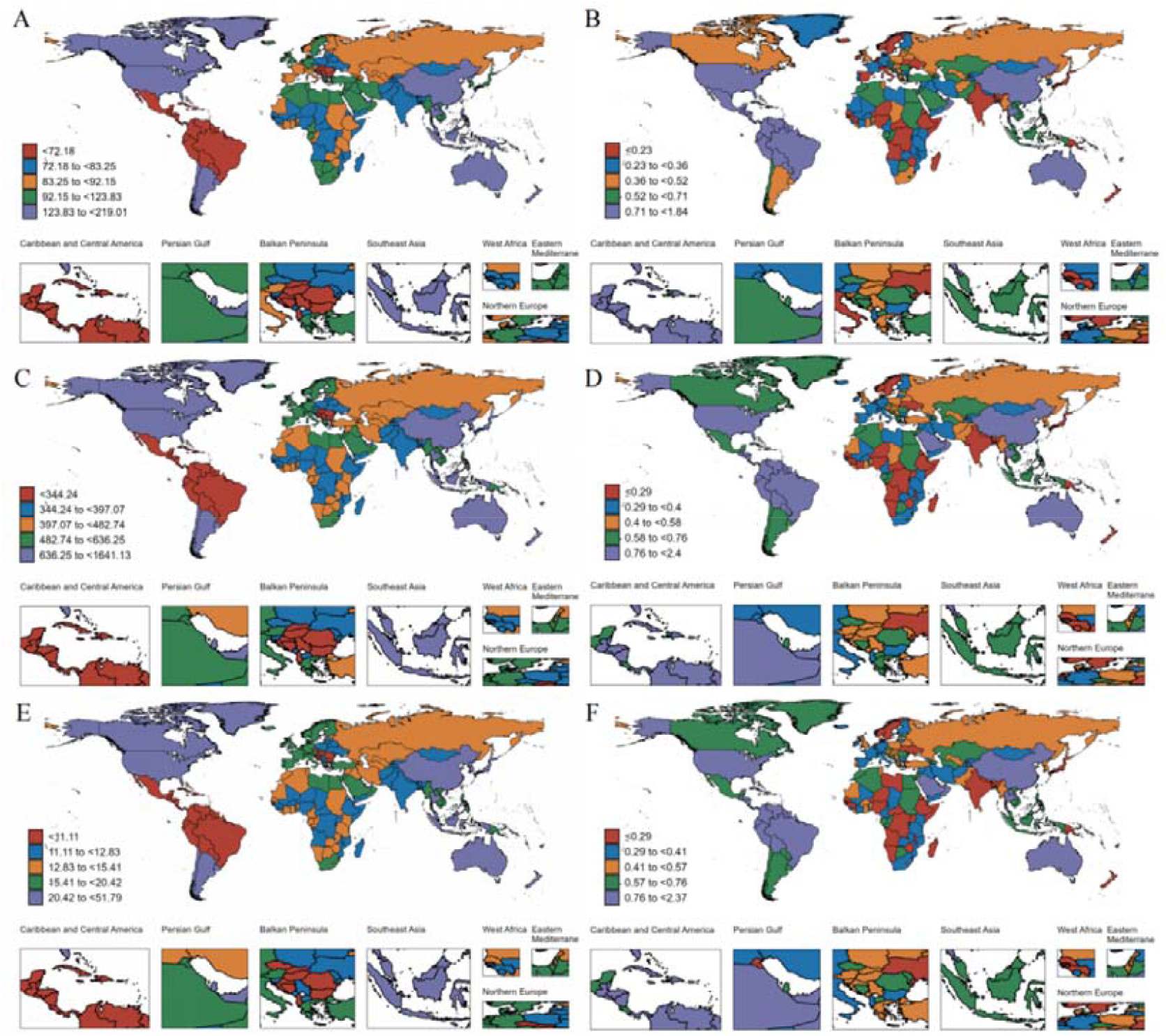
Global distribution of gout burden in 2021. (A) ASIR; (B)EAPC of incidence; (C) ASPR; (D) EAPC of prevalence; (E) ASYR; (F) EAPC of YLDs

The 2021 country-level data revealed a pronounced position correlation between the SDI and ASIR, ASPR, and ASYR for gout (Figure 5B, 5D, 5F). Specifically, countries with lower SDI, such as Haiti, Guatemala, and Honduras, had markedly lower-than-expected ASYR for these disease categories in 2021. In contrast, high-income nations such as the USA, Canada, and Greenland exhibited very high ASYR, corroborating the strong positive correlation between SDI and the burden of gout.

**Figure 5.**
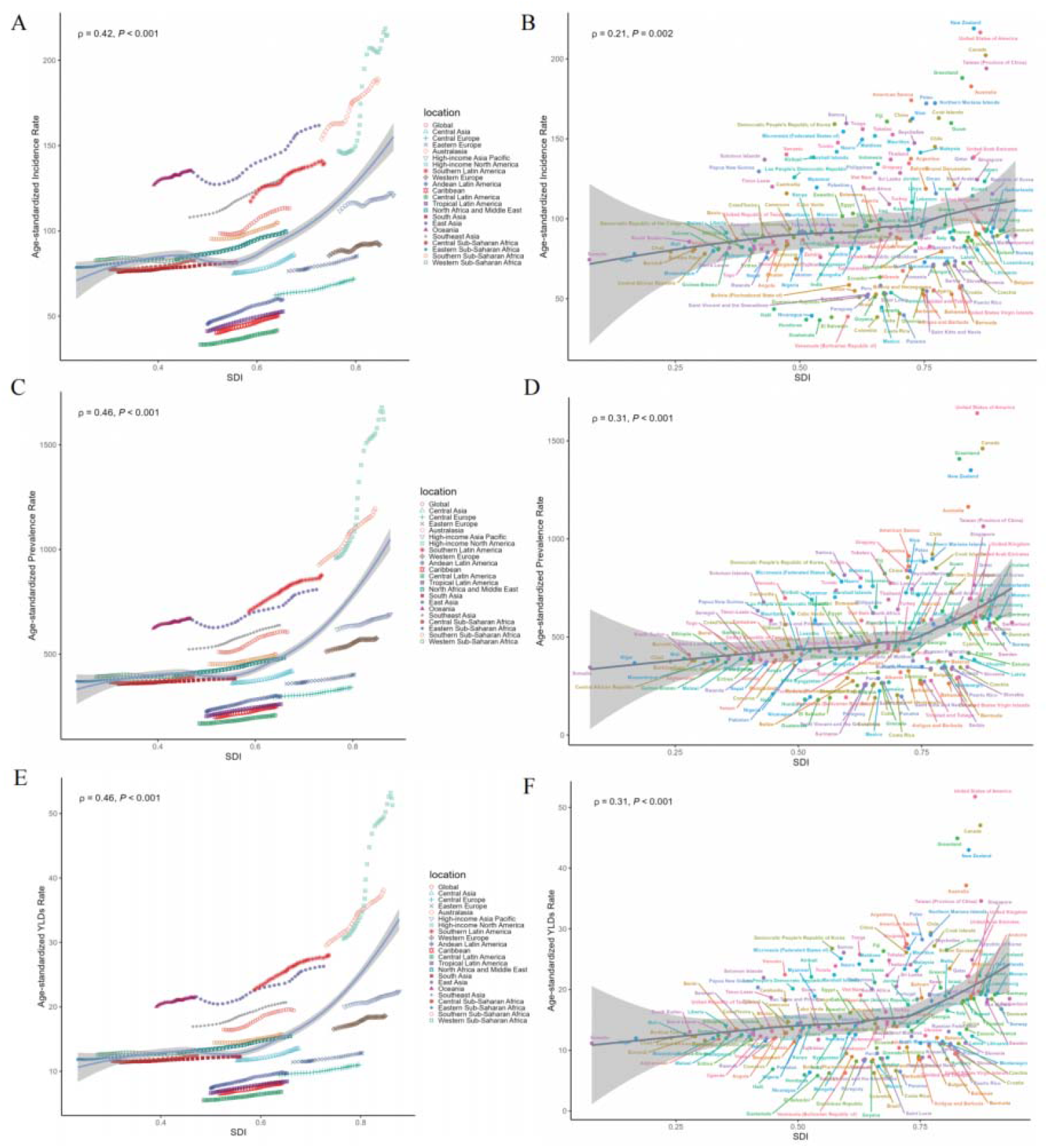
ASIR (A), ASPR (C) and ASYR (E) of gout globally, 21 GBD regions and ASIR (B), ASPR (D), and ASYR (F) of gout in 204 countries and territories by SDI In Figure 5, each data point represents the ASR and corresponding SDI value for a specific region and year. The thirty data points for each region depict the trends over the thirty years preceding and including the year 2021. The shaded area represents the 95% confidence interval (CI) of the expected values. Points above the solid line represent a higher-than-expected burden, and those below the line show a lower-than-expected burden.

### Burden of gout by SDI

Analysis of 21 GBD regions shows that as SDI levels rise, ASIR, ASPR, and ASYR all increase, with their growth rates accelerating with higher SDI levels. Among all 21 regions, central Latin America has the lowest ASYR. In high SDI regions, high-income North America has the highest ASYR, while Western Europe has the lowest ASYR. ASIR and ASPR show similar relationships with SDI (Figure 5A, 5C, 5E).

High levels of both absolute and relative SDI-related inequality in gout burden were observed. The slope index of inequality revealed an excess YLDs of 8.15 (95% CI, 6.10 to 10.21) per 100,000 in 1990 when comparing countries with the highest and lowest SDIs, while in 2021 this rose to 14.69 (95% CI, 11.90 to 17.48). The concentration index, which measures relative gradient inequality, also increased from a 1990 value of 0.16 (95% CI 0.13 to 0.19) to 0.28 (95% CI 0.25 to 0.32) in 2021, consistent with an unbalanced distribution of disease burden among countries with different socioeconomic development indices (SDIs). Higher SDI countries tended to shoulder a higher burden of gout. Analysis of the concentration index reveals that, from 1990 to 2021, the degree of concentration in China, the USA, and India has increased to varying degrees. In 2021, the degree of concentration in China and the USA was significantly higher than that in India (Table S3, Figure 6).

**Figure 6.**
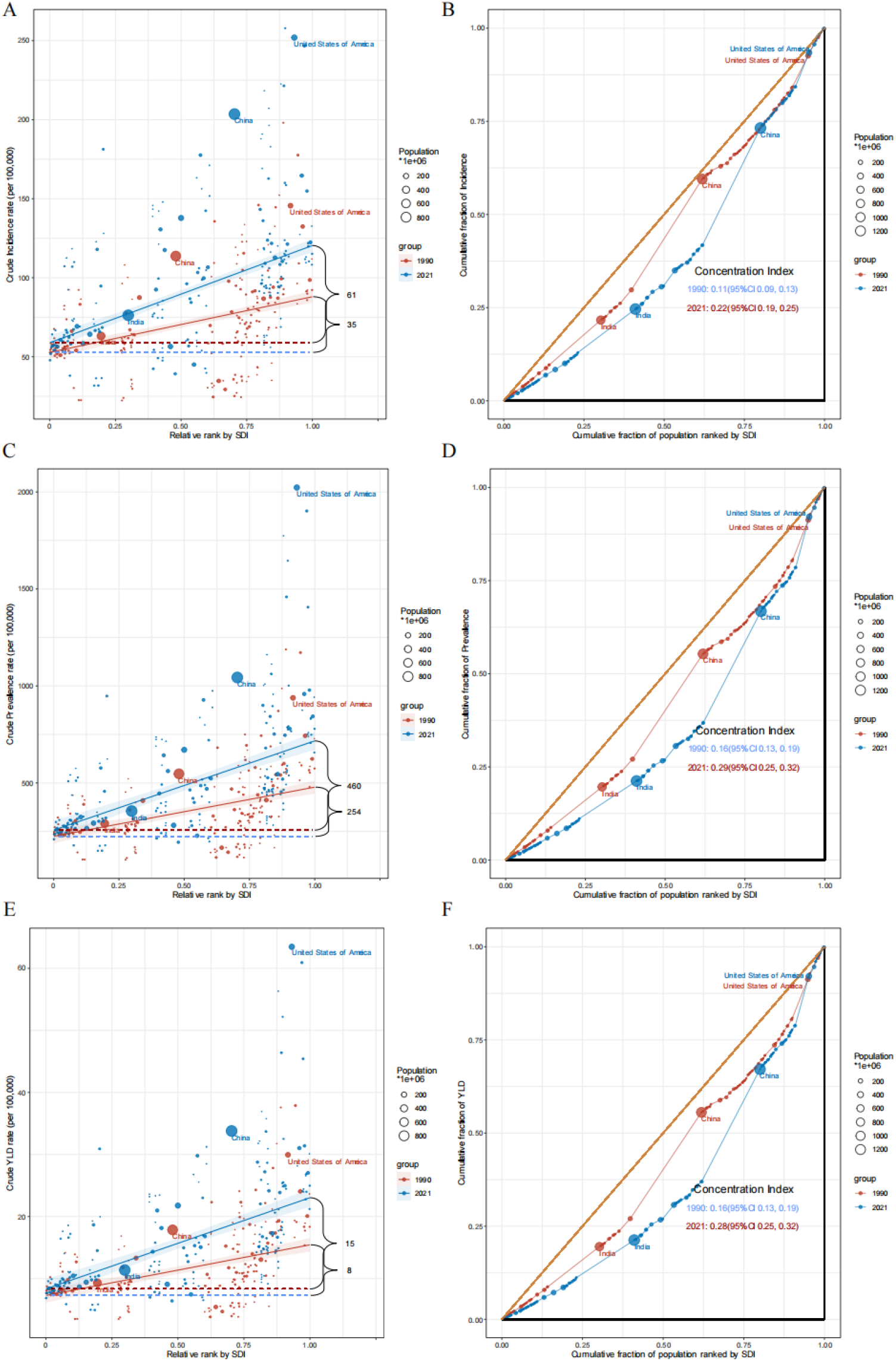
Wealth-based inequality of gout in populations living in countries with varying levels of socioeconomic development (SDI), 1990-2021. Slope index of inequality: absolute inequality of incidence (A), prevalence (C) and YLDs (E); Concentration curves of inequality: relative inequality of incidence (B), prevalence (D) and YLDs (F).

### Age and Sex Patterns

In 2021, ASIR, ASPR, and ASYR rates for gout in males were approximately four times those in females. Incident cases were estimated at 46.87 million in males versus 13.96 million in females; prevalent cases at 259.45 million versus 67.99 million; and YLDs at 8.34 million years versus 2.18 million years. Between 1990 and 2021, both sexes experienced a rising burden of gout. Although the EAPC in incidence was similar in males (0.62, 95% CI 0.57 to 0.67) and females (0.63, 95% CI 0.57 to 0.68), increases in prevalence (males 0.77 vs females 0.68) and YLDs (males 0.77 vs females 0.66) were higher in males than in females (Table S4).

We further stratified the analysis by 5-year age groups to assess the burden of gout by sex across different age intervals. Overall, both ASIR, ASPR, and ASYR rose with age in both sexes, and males had more than twice the burden of females in every age group. Among males, the most significant burden occurred at 55-59 years (5.31 million prevalent cases, 95% UI 3.70 to 7.21 million; 0.16 million YLDs, 95% UI 0.10 to 0.27 million years). In females, peak burden was at 60-64 years (5.31 million prevalent cases, 95% UI 3.70 to 7.21 million; 49,232.9 YLDs, 95% UI 27,598.9 to 80,078.4 years) (Table S5, Figure 7).

**Figure 7.**
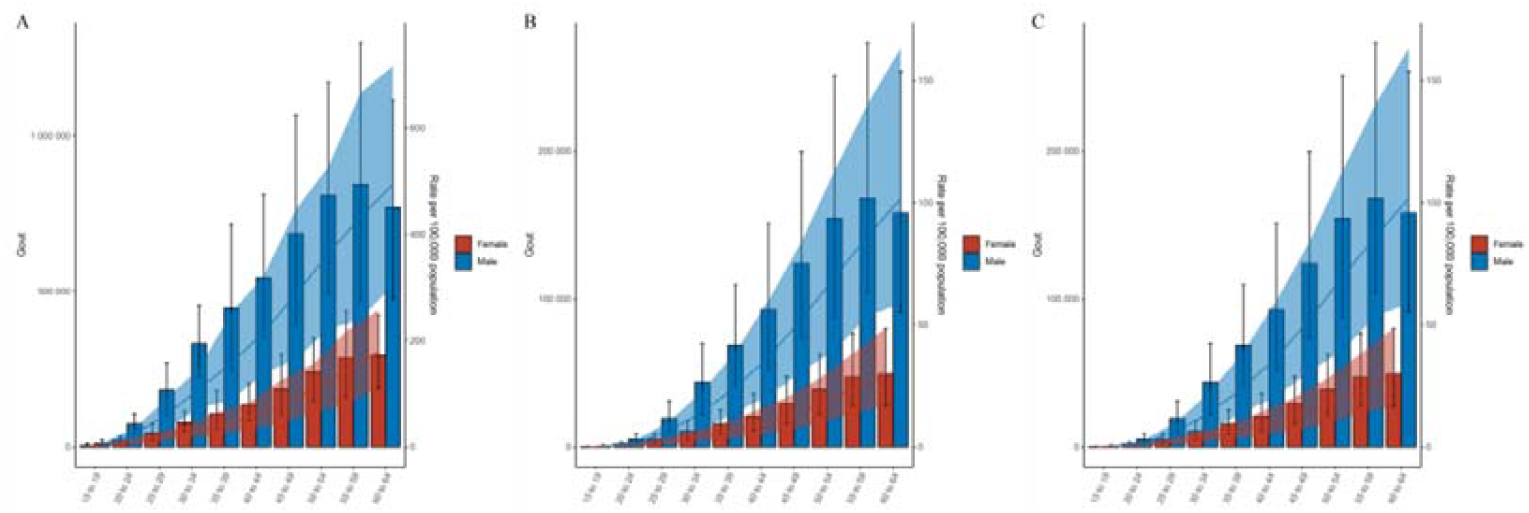
Number of incidence cases and ASIR (A), prevalence cases and ASPR (B), and YLDs and ASYR of gout in patients by gender and age Group, 2021.

### Decomposition analysis

From 1990 to 2021, YLDs due to gout among the working-age population increased substantially, with the most pronounced growth observed in middle-income development index (SDI) regions. Decomposition analysis of the total population revealed that population growth contributed 54.43% to the increase in YLDs, followed by population ageing (24.94%) and epidemiological change (20.63%). The relative contribution of these drivers varied by SDI level. In low, low-middle, and middle SDI regions, increases in YLDs were predominantly driven by population growth, accounting for 96.94%, 73.27%, and 48.61% of the total increase, respectively. This indicates that demographic expansion substantially intensified the burden of gout in these regions. With rising SDI levels, the contribution of epidemiological change became increasingly prominent, as follows: low SDI (7.00%), low-middle SDI (11.55%), middle SDI (14.05%), high-middle SDI (32.09%), and high SDI (47.00%). Notably, in low SDI regions, ageing exerted a negative contribution (-3.94%) to changes in YLDs, suggesting that population ageing in these settings may have attenuated the gout burden (Figure 8).

**Figure 8.**
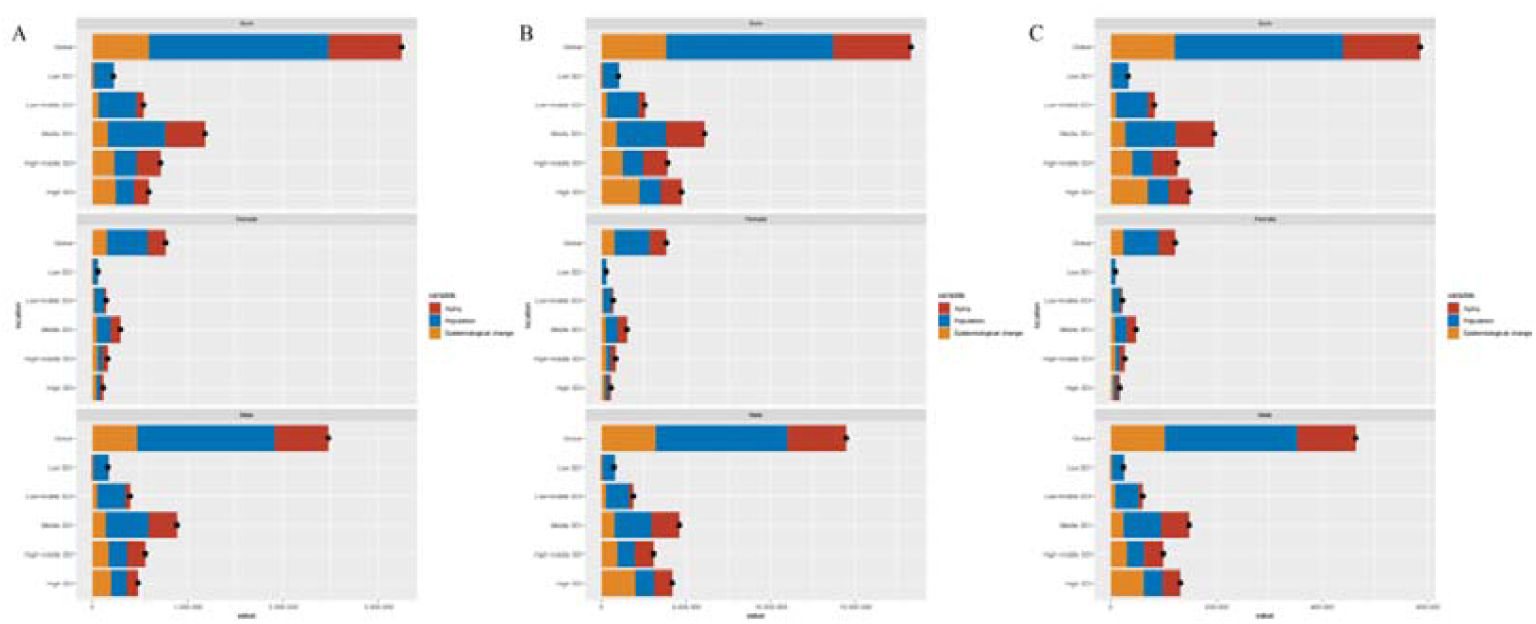
Decomposition analysis of YLD changes in gout according to population-level determinants from 1990 to 2021 at the global level and by SDI.

Sex-specific decomposition analyses showed that the pattern of change in YLDs among males closely mirrored that of the total population. At the same time, distinct trends were observed among females across SDI strata. YLDs in working-age females increased with SDI to a peak in middle-SDI regions before declining at higher SDI levels. In high SDI regions, unlike in males, where the increase in YLDs was mainly driven by epidemiological change, the contributions of ageing (37.83%), population growth (32.43%), and epidemiological change (29.74%) to the YLD increase in females were relatively balanced. These findings highlight the significant role of epidemiological changes in contributing to the increased disease burden among working-age males (Figure 8).

## Discussion

In this comprehensive analysis of gout in the working-age population, we found that gout poses a substantial and growing global health burden among individuals aged 15–64 years. As of 2021, over 32 million working-age adults worldwide are affected by gout, and that number is projected to climb to ∼45 million by 2045. Gout incidence and prevalence in this demographic have risen steadily since 1990 (EAPC around +0.7–0.8% per year for age-standardised rates), mirroring global trends observed in all-age analyses, though at a slightly tempered pace(Li et al., 2025, Li et al., 2024). The burden is unevenly distributed: high-income regions and males bear a disproportionately large share of gout cases, and inequalities between countries have widened over the past three decades. Our decomposition of trends indicates that population expansion and ageing are the major drivers of the increasing gout caseload, while a smaller yet significant portion of the rise is due to actual increases in disease occurrence (likely tied to lifestyle changes). Importantly, this study is the first to specifically focus on gout in the working-age group at a global level, revealing that a majority of gout patients are under 65, in their prime working years, which has implications for clinical management and policy prioritisation.

Our findings are broadly consistent with previous GBD-based studies on gout but provide new insights specific to working-age adults. The GBD 2021 gout burden analysis PMID: 38996590IF: 15.0 Q1 by Chen *et al*. (GBD Collaborators) reported 55.8 million gout cases globally (all ages) in 2020 and highlighted a 22.5% increase in age-standardised prevalence since 1990(2024c). We observed a similar trend in working-age individuals, though the increase was more modest (∼20%), as older populations have experienced sharper rises. For instance, Li *et al*. (2024) examined gout in adults aged≥55 and found an EAPC of +1.08 % per year in prevalence.(2024c) – higher than our ∼0.8%/year for <65 – indicating the epidemic of gout has been more pronounced among older adults. With ∼32.7 million gout cases in the 15–64 group, our study confirms that a substantial portion of global burden lies within the working-age population.

Sex differences in gout burden also mirrored previous reports. We found a global male-to-female prevalence ratio of ∼3:1 in working-age individuals, consistent with epidemiological data all-age estimates (2024c). However, our data hint at a narrowing gap, with mid-life female gout rates rising—possibly due to obesity and improved detection.

Geographically, our findings reaffirm that high-income regions and Pacific nations show the highest gout rates. The Australasia region (Australia and New Zealand) consistently emerges as having among the highest gout rates (Zhang et al., 2025), which we also observed for working-age adults. Prior literature attributes this to genetic predispositions (e.g., prevalence of risk alleles in Maori and Pacific Islander populations) and high rates of obesity and comorbidities in these countries. High-income North America and parts of Oceania likewise have very high gout burdens (Zhang et al., 2025). Conversely, sub-Saharan Africa and some Asian regions exhibit lower rates, possibly reflecting historically less obesogenic diets and underdiagnosis. Our work specifically extended prior findings by quantifying the inequality metrics for working-age adults, showing that both absolute and relative disparities in gout burden have widened over time, with higher SDI countries increasingly concentrated at the top.

Compared to Xia *et al*. (2023), who an examined younger adults (15–39 years), we show that most of the burden increase is concentrated in ages 40–64.Gout incidence rises with age, our results indicate that peak gout burden occurs in the later working years (50s and early 60s), a critical period for workforce participation and economic productivity.

A key novel contribution of our study is the decomposition analysis for the working-age gout trends. We quantified drivers of burden increase from 1990 to 2021: ∼54% was due to population growth, ∼25% to rising rates, and ∼20% to ageing. These results align with global trends(Han et al., 2024) but provide more detailed insights by SDI and sex. In high-income settings, epidemiological changes contributed a larger share to prevalence growth, reflecting rising obesity and metabolic syndrome (2024c).

Our projections indicate that, while absolute case numbers will continue rising, age-standardised rates in working-age adults are likely to stabilise. This contrasts with steeper projected growth in older populations (Wang et al., 2025). Assuming stable risk factor trends and improved management, gout incidence may plateau—offering an opportunity for intervention before the burden escalates further.

The unique focus of this study on the working-age population reveals essential insights. First, we demonstrated that over half of global gout cases now occur before age 65. This highlights gout as a condition that significantly affects people in their productive years, not just retirees. The implications for health systems and economies are noteworthy: gout flares cause pain and disability that can reduce work productivity and increase absenteeism.(Sigurdardottir et al., 2018). Chronic uncontrolled gout can lead to joint damage and disability, potentially even causing early retirement in severe cases. Therefore, gout in mid-life represents not only a healthcare burden but also an economic burden due to lost productivity. Our findings should alert policymakers that addressing gout is part of maintaining a healthy workforce.

Second, by isolating this age group, we can appreciate the opportunity for prevention and early intervention. Many risk factors for gout (obesity, diet, and alcohol use) often become entrenched in young adulthood and mid-life. Tackling these factors in the working-age population could yield dual benefits – improving immediate health and preventing progression to severe gout and comorbidities in older age. For example, weight management and dietary modification in a 40-year-old with hyperuricaemia could prevent them from developing tophaceous gout or severe joint destruction by age 70. Our study highlights the importance of public health initiatives targeting lifestyle risk factors among younger adults, not just older individuals, to effectively reduce the future gout burden.

Third, our analysis of inequalities, specifically in the working-age population, adds nuance to global health equity discussions. We found widening disparities with a higher burden in developed regions. This paradox, where wealthier societies have more gout, likely relates to lifestyle “luxury” factors (high-calorie diets, sedentary jobs) as well as possibly better detection. It suggests that as low-income countries develop economically, they may face a surge in gout unless preventive measures are in place (similar to trends seen with diabetes and other lifestyle diseases). Thus, there is a window now for low- and middle-income countries to implement strategies (like promoting healthy diets, regulating sugary beverage and alcohol consumption, and encouraging physical activity) to avoid repeating the high-SDI countries’ trajectory of gout escalation.

From a clinical perspective, focusing on working-age gout patients raises considerations about management strategies tailored to this group. Working-age patients may have different treatment challenges compared to older patients – they might be less adherent to daily urate-lowering therapy due to busy work/family life, they may prioritise acute relief to minimise work disruption, and they often face different comorbidities (e.g., more metabolic syndrome, less renal impairment than older cohorts). Ensuring that primary care and occupational health systems screen for hyperuricaemia in high-risk individuals (like those with obesity or heavy alcohol use) and manage gout aggressively could prevent long-term complications. Additionally, given the stigma and trivialisation that sometimes surround gout (as a self-inflicted disease of gluttony), education campaigns in workplaces could help improve understanding that gout is a biological illness that can be effectively treated, encouraging patients to seek care rather than silently endure flares.

This study offers the first comprehensive assessment of gout burden among the global working-age population (15–64 years) using GBD 2021 data across 204 countries over three decades. By applying a robust analytical methods – including EAPC trend analysis, joinpoint regression, inequality indices, and decomposition we quantified the contributions of population growth, ageing, and epidemiologic shifts to rising gout burden, and provided projections to 2045. These insights complement existing studies on older adults and enhance understanding of modifiable versus demographic drivers.

However, several limitations warrant caution. Our estimates depend on GBD modelling, which may be affected by limited primary data in low-SDI countries, potentially underestimating true burden and inflating observed inequalities. The GBD framework does not distinguish first-ever from chronic gout or capture age of onset and severity variation, limiting insights into changing disease patterns. Disability weights assume average severity, potentially masking the disproportionate impact on certain subgroups, such as manual labourers. Forecasts are based on historical trends and may not reflect future changes in risk factor trajectories.

Lastly, while our focus on the 15–64 age group aligns with standard definitions, we acknowledge that gout burden continues beyond this threshold and future studies should consider the full age continuum.

Our findings have several important implications for both public health policy and clinical care. First, gout prevention should be explicitly integrated into broader non-communicable disease (NCD) strategies. The strong association between gout and modifiable risk factors—particularly elevated body mass index (BMI), poor diet, and alcohol use—suggests that interventions designed to address cardiovascular disease, diabetes, and obesity will also be effective in reducing the burden of gout (2024c). Public health campaigns focusing on dietary education, salt and sugar reduction, physical activity promotion, and healthy weight maintenance in high-risk regions (such as the Pacific Islands and urbanised parts of Asia) could yield long-term reductions in hyperuricaemia and incident gout. Our results underscore the value of targeting these efforts toward adults in their 20s, 30s, and 40s—before gout onset or early in its course—when lifestyle changes can significantly reduce cumulative disease burden.

Second, the growing burden of gout in working-age adults warrants greater recognition of gout in workplace health policies. Employers and occupational health services should acknowledge that gout can be a cause of intermittent but significant disability, particularly in middle-aged employees. Supportive workplace strategies such as flexible leave policies during acute flares, access to hydration and healthy food options, and insurance coverage for long-term gout management could help minimise productivity losses. Educational efforts in the workplace should also help dispel common misconceptions, such as the belief that gout is solely due to indulgent lifestyles, and promote timely medical management.

Third, the healthcare system must strengthen the delivery of gout care, especially within primary care settings where most working-age patients are first diagnosed. Despite the availability of effective urate-lowering therapies such as allopurinol and febuxostat, treatment is often suboptimal in routine practice. Physicians should proactively initiate and up-titrate urate-lowering medication in patients with recurrent flares and monitor serum urate to achieve therapeutic targets. Given that younger patients may discontinue medication after symptom improvement, there is a need for structured patient education, follow-up, and adherence support. Gout should be managed as a chronic disease—akin to hypertension or type 2 diabetes—requiring long-term maintenance to prevent cumulative joint damage and disability.

Fourth, our findings highlight increasing global inequalities in gout burden and the need for international collaboration to address them. High-income countries, which currently bear the highest burden, can support lower-income settings through data sharing, training initiatives, and access to essential diagnostics and medications. Strengthening clinical capacity in low-resource regions—for example, through training in joint aspiration, ultrasound techniques, and diagnostic criteria—would improve early detection and treatment. Ensuring the availability and affordability of key drugs such as colchicine, NSAIDs, and urate-lowering therapies in low-income countries is a crucial step toward equitable gout care. Conversely, understanding how some countries maintain relatively low gout rates—possibly through diet patterns, public health infrastructure, or genetic differences—may provide valuable insights for prevention in regions with higher gout burdens.

Ultimately, this study highlights several key priorities for future research. More granular data on the age of gout onset and longitudinal disease progression are needed, as the GBD framework does not currently capture the age at first attack or severity levels. Prospective cohort studies tracking early onset and chronic progression in different populations would be valuable for risk stratification. Research focusing on female gout is also overdue, as sex-specific clinical presentations and risk profiles remain underexplored. Implementation research is urgently needed to evaluate the most effective way to integrate gout management into primary care, particularly in low- and middle-income settings. In addition, given the central role of obesity in gout pathogenesis, future work on effective obesity prevention, including behavioural, public health, and pharmacologic interventions, may offer indirect but consequential avenues for gout control at scale.

## Conclusion

In conclusion, this study demonstrates that gout is a widespread and rising health issue among the global working-age population, with tens of millions affected and more to come as populations grow and age. Far from being solely a disease of retirees, gout affects many individuals in their most productive years, underscoring the need for age-targeted prevention and management. Demographic trends primarily fuel the growing burden in mid-life. Still, they are also accentuated by lifestyle factors – a reminder that addressing modifiable risks like obesity, diet, and alcohol use is crucial. We observed widening disparities, with higher-income regions currently facing the most significant gout burden, but lower-income countries likely to catch up as lifestyles change. To avert a future crisis of gout-related disability, concerted action is needed now. Public health policies must integrate gout and hyperuricaemia control into broader non-communicable disease (NCD) prevention strategies, and healthcare systems should strengthen early detection and long-term care for gout patients. By doing so, we can reduce the pain and disability from gout in the working population, improve quality of life, and maintain economic productivity. Gout is an ancient disease, but its escalating impact in today’s working adults is a modern warning sign – one that we ignore at our peril. Effective, age-tailored interventions implemented today will pay dividends in curbing the global gout burden of tomorrow.

## Data Availability

All data produced in the present study are available upon reasonable request to the authors

## Supplement figures

**Figure S1.**
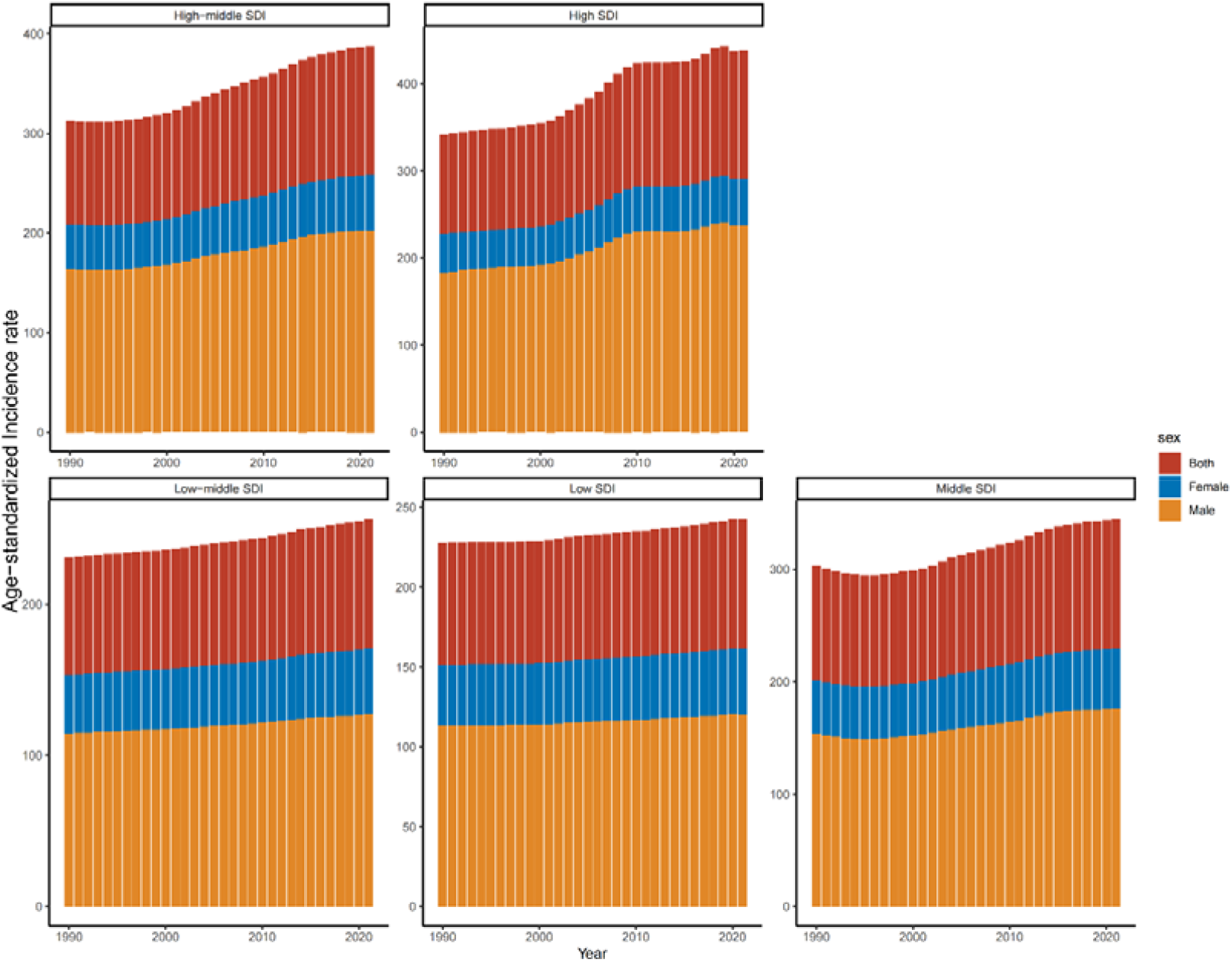

**Figure S2.**
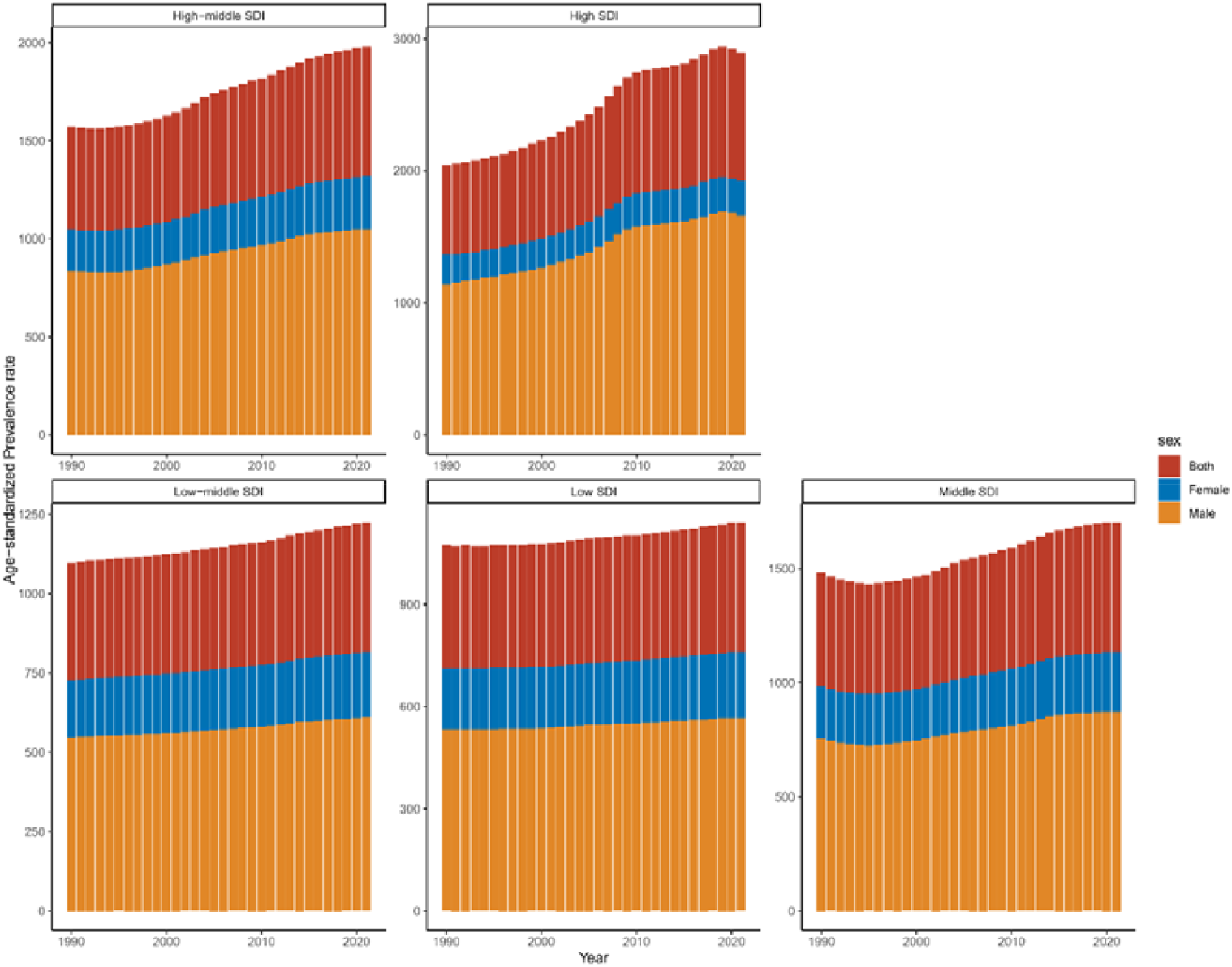

**Figure S3.**
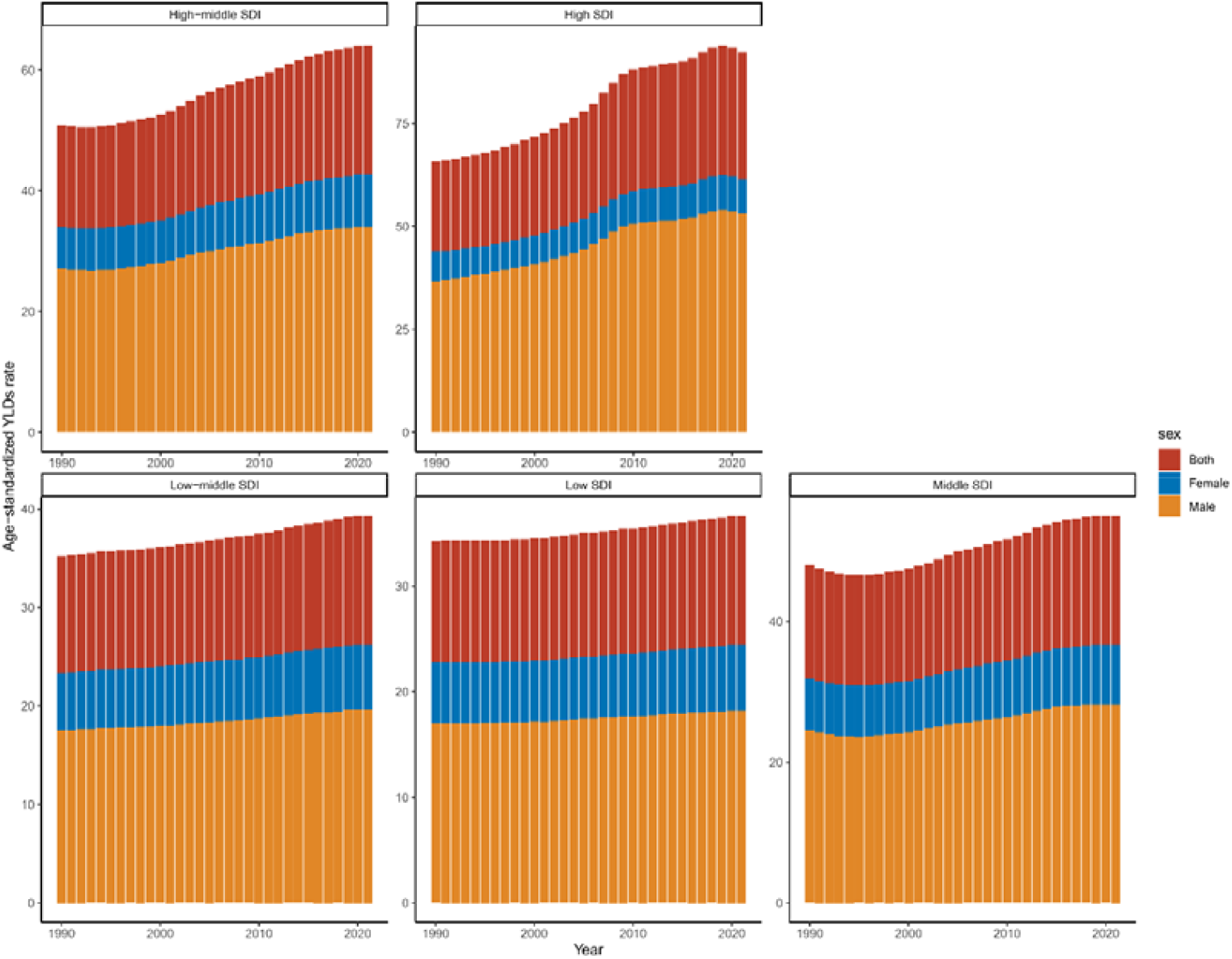

## References

.Available: https://vizhub.healthdata.org/gbd-results/ [Accessed].

2024a. Burden of disease scenarios for 204 countries and territories, 2022-2050: a forecasting analysis for the Global Burden of Disease Study 2021. Lancet, 403, 2204–2256.

2024b. Global incidence, prevalence, years lived with disability (YLDs), disability-adjusted life-years (DALYs), and healthy life expectancy (HALE) for 371 diseases and injuries in 204 countries and territories and 811 subnational locations, 1990-2021: a systematic analysis for the Global Burden of Disease Study 2021. Lancet, 403, 2133–2161.

2024c. Global, regional, and national burden of gout, 1990-2020, and projections to 2050: a systematic analysis of the Global Burden of Disease Study 2021. Lancet Rheumatol, 6, e507–e517.

2025. Gout. Annals of Internal Medicine, 178, ITC33–ITC48.

Dalbeth, N., Gosling, A. L., Gaffo, A. & Abhishek, A. 2021a. Gout. Lancet, 397, 1843–1855.

Dalbeth, N., Gosling, A. L., Gaffo, A. & Abhishek, A. 2021b. Gout. The Lancet, 397, 1843–1855.

Danve, A. & Neogi, T. 2020. Rising Global Burden of Gout: Time to Act. Arthritis Rheumatol, 72, 1786–1788.

Dehlin, M., Jacobsson, L. & Roddy, E. 2020. Global epidemiology of gout: prevalence, incidence, treatment patterns and risk factors. Nature Reviews Rheumatology, 16, 380–390.

Han, T., Chen, W., Qiu, X. & Wang, W. 2024. Epidemiology of gout - Global burden of disease research from 1990 to 2019 and future trend predictions. Ther Adv Endocrinol Metab, 15, 20420188241227295.

Hankey, B. F., Ries, L. A., Kosary, C. L., Feuer, E. J., Merrill, R. M., Clegg, L. X. & Edwards, B. K. 2000. Partitioning linear trends in age-adjusted rates. Cancer Causes Control, 11, 31–5.

Hosseinpoor, A. R., Bergen, N., Kirkby, K. & Schlotheuber, A. 2023. Strengthening and expanding health inequality monitoring for the advancement of health equity: a review of WHO resources and contributions. Int J Equity Health, 22, 49.

Kim, H. J., Fay, M. P., Feuer, E. J. & Midthune, D. N. 2000. Permutation tests for joinpoint regression with applications to cancer rates. Stat Med, 19, 335–51.

Kleinman, N. L., Brook, R. A., Patel, P. A., Melkonian, A. K., Brizee, T. J., Smeeding, J. E. & Joseph-Ridge, N. 2007. The impact of gout on work absence and productivity. Value Health, 10, 231–7.

Li, M., Nie, Q., Xia, Q. & Jiang, Z. 2025. Assessing cross-national inequalities and predictive trends in gout burden: a global perspective (1990-2021). Front Med (Lausanne), 12, 1527716.

Li, Y., Chen, Z., Xu, B., Wu, G., Yuan, Q., Xue, X., Wu, Y., Huang, Y. & Mo, S. 2024. Global, regional, and national burden of gout in elderly 1990–2021: an analysis for the global burden of disease study 2021. BMC Public Health, 24, 3298.

Sigurdardottir, V., Drivelegka, P., SvÄRd, A., Jacobsson, L. T. H. & Dehlin, M. 2018. Work disability in gout: a population-based case-control study. Ann Rheum Dis, 77, 399–404.

Wang, J., Zhao, Y., Zhao, B. & Zhang, Y. 2025. Global burden and future projections of geriatric gout (1990-2021): a comprehensive analysis and Bayesian Age-Period-Cohort modeling. Front Public Health, 13, 1577265.

Zhang, Y., Jin, Z., Yao, J., Wang, D., Yunxian, Y. & Zhang, W. 2025. Global, regional, and national burden of gout in people aged 15-39 years from 1990 to 2021: trends, cross-country inequalities and forecast to 2035. Joint Bone Spine, 105929.

